# Association of the *FTO* rs9939609 variant with glycemic control

**DOI:** 10.64898/2026.03.05.26347689

**Authors:** Nicolas Fragoso-Bargas, Rocío Vanessa Escarcega-Castro, Irma Quintal-Ortiz, Ligia Vera Gamboa, Guillermo Valencia-Pacheco, Nina Valadez-González

## Abstract

Type 2 diabetes (T2D) affects 11.1% of the global population, underscoring the need for biomarkers that inform treatment response and glycemic outcomes. We evaluated the association between the *FTO* variant rs9939609-A and glycemic control in a Mexican population. A total of 174 individuals living with T2D from Mérida and Sisal, Yucatán, were included, of whom 85% were receiving oral hypoglycemic agents as main treatment. Glycemic control was defined cross-sectionally as good (≤130 mg/dL, n=63) or poor (>130 mg/dL, n= 111) with fasting glucose. Linear mixed models incorporating relevant covariates and a family random intercept were used. Effect size estimates were transformed to logit odds ratios. After adjustment for age, sex, BMI, years with T2D, and treatment, we observed a significant association in the additive (OR = 1.15 [1.003–1.31]) and recessive (OR = 1.51 [1.03–2.23]) models. To conclude, rs9939609-A may be associated with poorer glycemic control despite pharmacologic therapy.

## Introduction

Type 2 diabetes (T2D) is a multifactorial disease characterized primarily by insulin resistance, followed by gradual dysfunction of pancreatic β-cells, which together lead to chronic hyperglycemia (1). The global prevalence of diabetes is 11.1%, and this number is projected to increase by approximately 45% by 2050 (2), i.e., to about 16.1%. This growing burden highlights the need to better understand the biological determinants of glycemic control, including genetic factors that influence treatment response and optimal glucose regulation.

Diabetes is generally managed with diet and life-style changes and, if necessary, with oral hypoglycemic agents (OHA) such as metformin and sulfonylureas (3). Treatment is considered successful when the patient achieves adequate glycemic control (4). According to standard clinical definitions, good glycemic control includes a fasting plasma glucose ≤130 mg/dL (5). When adequate glycemic control cannot be achieved with OHA treatment, insulin therapy, whether as monotherapy or as an adjunct to OHA treatment, may be considered (6). Regardless of the treatment modality, failure to meet the recommended glycemic targets is considered poor glycemic control (7).

Thus far, eight GWAS have been conducted to identify genetic factors associated with treatment response in T2D (8–15), but the number of associated loci remains very small compared with those identified for T2D diagnosis. While T2D GWAS have identified ∼1,300 loci using sample sizes of approximately 2.5 million individuals (16), GWAS on glycemic control have been considerably underpowered, including only ∼300–4,000 participants, limiting discovery. As a result, only around 10 SNVs have been robustly associated with glycemic control and related phenotypes (8,11–15). Consequently, the genetics underlying glycemic control and treatment response in T2D remain far less explored than the genetic architecture of T2D onset itself. In the absence of well-powered GWAS, examining candidate genes within previously validated T2D loci may represent an immediate and pragmatic strategy for identifying variants associated with glycemic control.

The fat mass and obesity-associated gene (*FTO*) spans approximately 400 kb and is located on chromosome 16q12.2 (17). One of its most widely studied genetic variants, rs9939609-A, has been consistently associated with body mass index (BMI) and obesity across diverse populations (18,19). However, rs9939609-A has also shown consistent associations with T2D (20), and in Northern European (21), Vietnamese (22) and Southeastern Mexico (23) populations, this association appears to be independent of BMI. In the same Southeastern Mexican population, where Mayan ancestry is prevalent, rs9939609-A was also associated with fasting glucose levels (23), further supporting a potential role for *FTO* in glycemic regulation. Given its well-documented associations with T2D risk and fasting glucose, rs9939609-A represents a strong biological candidate for contributing to glycemic control among individuals with established T2D.

Therefore, the aim of this study is to assess cross-sectional associations between rs9939609-A and clinically defined glycemic control (fasting plasma glucose ≤130 mg/dl) in a well-characterized Mexican population, leveraging detailed data on diabetes treatment and clinical management.

## Materials and methods

### Study population

For this study, we included patients with T2D from two populations in Yucatan, Mexico. The first study was a case-control study conducted in the locality of San Jose Tecoh in Merida, Yucatan (23). All participants were recruited at the health center “Unidad Universitaria de Inserción Social (UUIS) San José Tecoh” between 2018 and 2019. Cases were included if they had a diagnosis of T2D according to Mexican guidelines (NOM-015-SSA2-2010), defined as fasting plasma glucose ≥126 mg/dL, random glucose ≥200 mg/dL, or ≥200 mg/dL two hours after a 75-g oral glucose load; were over 20 years old; were of any sex; and were born in Mérida, Yucatán. Controls were required to have no family history of T2D for at least two generations; be over 20 years old; be of any sex; and be born in Mérida, Yucatán. General exclusion criteria included being pregnant, having a relative already enrolled in the study, and, in the case group, having type 1 diabetes. A total of 92 cases and 92 controls were recruited and had complete genotyping data.

The second population comes from a population-based study conducted in Sisal, Yucatán, México, between 2010 and 2012. Sisal is a small coastal community with a strong Mayan cultural heritage (24). Individuals were eligible if they were born in Sisal and had deep local family roots, defined as having relatives who had lived in the town for at least three generations, as verified through detailed family information. Recruitment focused on individuals with T2D and their metabolically healthy relatives, and only participants aged 20 years or older were included. Exclusion criteria included pregnancy and diagnoses of diabetes types other than T2D. In total, 106 individuals with T2D and 145 without T2D were enrolled. Of all participants, 99 individuals with T2D and 133 without T2D were successfully genotyped and had complete age information.

From the 92 individuals with T2D from San Jose Tecoh and the 99 from Sisal, only those with complete clinical information relevant to this study and complete genotyping were included. Four participants were excluded because their fasting glucose values were below 70 mg/dL. The final sample consisted of 81 participants from San Jose Tecoh and 93 from Sisal, giving a total of 174 individuals.

All participants from both study populations provided written informed consent, confirming their voluntary participation in accordance with the Declaration of Helsinki and the Mexican General Health Law. The study protocol was reviewed and approved by the Ethics Committee of the Dr. Hideyo Noguchi Regional Research Center at the Autonomous University of Yucatán (UADY). The approval code was CEI-CIR-UADY-007-2011, and the approval date 13.5.2011.

### Anthropometrics and biochemical clinical measurements

Age, years living with T2D, height, and weight were recorded at recruitment in both study populations, and body mass index (BMI) was calculated from these measurements. Fasting plasma glucose was measured in all participants using the same Roche Cobas c111 analyzer (Roche Diagnostics Ltd., Rotkreuz, Switzerland) following the manufacturer’s guidelines.

### Cross-sectional determination of glycemic control categories

Glycemic control was assessed according to the Mexican NOM 015 SSA2 2010 clinical criteria, which, amongst its definitions, determines good control as a fasting plasma glucose ≤130 mg/dL and poor control as values at or above this threshold. Based on this criterion, 111 participants were classified as having poor glycemic control and 63 as having good control.

### Genotyping

DNA was extracted from peripheral blood using the Macherey-Nagel® kit (Macherey-Nagel GmbH & Co. KG, Düren, Germany) following the manufacturer’s protocol. Real-time PCR genotyping of the rs9939609 variant was performed in all participants using the ECO™ Illumina® platform. A TaqMan® allele discrimination assay with the specific probe C 30090620_10 (Applied Biosystems, Foster City, CA, USA) was used, following the manufacturer’s recommended protocol. Each run included positive and negative controls.

### Statistical analyses

Hardy-Weinberg equilibrium was assessed in the full sample and in the poor and good control groups. We applied the Exact test described by Wigginton and collaborators (25), which provides appropriate control of type I error in both large and small datasets. All calculations were performed using the Hardy-Weinberg R package (26–28) version 1.7.9. Association analyses were performed under the following genetic models: additive (TT = 2, AT = 1, AA = 0), dominant (TT or AT = 1, AA = 0), overdominant (AT = 1, TT or AA = 0), and recessive (AA = 1, TT or AT = 0). For all analyses, the minor T allele was considered the risk allele. In San Jose Tecoh, all individuals were unrelated, whereas in Sisal there were several families. Although most participants were unrelated (i.e., singletons), we accounted for relatedness to avoid residual confounding. To do so, we used mixed linear regression with family as a random intercept in all models. Although mixed logistic regression is theoretically more suitable for binary outcomes, it often fails to converge when genotype groups are small, particularly in recessive models. Linear mixed models tolerate sparse genotype categories better and are widely used in GWAS for binary traits such as T2D (29). For consistency across analyses, we therefore used linear mixed models and exponentiated the coefficients and confidence intervals to approximate effects on the odds scale.

We constructed the following models: 1) crude model without adjustment, 2) adjusted for age and sex, 3) adjusted for age, sex, years with T2D, and treatment, and 4) the same as model 3 with the addition of BMI. All models accounted for relatedness as described above. For the treatment variable, we had seven schemes: 1) only diet control, 2) metformin, 3) glibenclamide, 4) metformin and glibenclamide, 5) pioglitazone, 6) insulin monotherapy, and 7) insulin with metformin. Because sample sizes varied widely across these schemes and including all categories would likely collapse or overfit the models, we collapsed treatment into three broader categories: 1) only diet control, 2) OHAs, and 3) insulin use. These three categories were used for modeling. We considered p<0.05 statistically significant and p<0.1 a statistical trend or marginal association. All models were constructed in R Studio (version 2025.09.1+401).

## Results

### Population characteristics

Table 1 presents the population characteristics. Of the 174 individuals included in this study, 111 were classified in the poor control group and 63 in the good control group. The mean age of the population was 57.29 years, with individuals in the poor control group averaging 55.95 years and those in the good control group averaging 59.65 years. The vast majority (85%) of the population were treated with OHAs alone. We had a total of 146 families, of which 127 were singletons. In the poor control group, we observed 99 families, of which 91 were singletons, and in the good control group there were 59 families, of which 55 were singletons. Approximately 68% of individuals were female. No individuals included in this study had missing values.

**Table 1:** Clinical and demographic characteristics of the studied population. Treatment scheme supergroups are shown in bold, followed by their specific subcategories.

| Phenotype | N | All (n=174) | N | Poor control<br>(n=111) | N | Good control<br>(n=63) |
| --- | --- | --- | --- | --- | --- | --- |
| Age | 174 | 57.29 (11.66) | 111 | 55.95 (11.76) | 63 | 59.65 (11.20) |
| Female sex (% of all studied population) | 119 | 119 (68.39) | 74 | 74 (67) | 45 | 45 (71.43) |
| Family (median number of members [range])* | 146 | 1[1–5] | 99 | 1[1-4] | 59 | 1 [1-2] |
| BMI (kg/m<sup>2</sup>) | 174 | 32.05 (6.96) | 111 | 32.76 (7.15) | 63 | 30.80 (6.48) |
| Fasting glucose (mg/dL) | 174 | 168.30 (64.81) | 111 | 173.37 (71.47) | 63 | 108.56 (15.40) |
| Years with T2D | 174 | 9.90 (7.62) | 111 | 9.93 (7.52) | 63 | 9.85 (7.87) |
| **Only diet control** | 11 | 11 (6.32) | 9 | 9 (8.11) | 2 | 2 (3.18) |
| **Oral hypoglycemics** | 148 | 148 (85.06) | 90 | 90 (81.10) | 58 | 58 (92.06) |
| Metformin | 42 | 42 (24.14) | 21 | 21 (18.92) | 21 | 21 (33.33) |
| Glibenclamide | 25 | 25 (14.37) | 13 | 13 (11.71) | 12 | 12 (19.05) |
| Glibenclamide and Metformin | 80 | 80 (45.98) | 56 | 65 (50.45) | 24 | 24 (38.10) |
| Pioglitazone | 1 | 1 (0.57) | 0 | 0 (0) | 1 | 1 (1.59) |
| **Insulin use** | 15 | 15 (8.62) | 12 | 12 (10.81) | 3 | 3 (4.48) |
| Insulin monotherapy | 7 | 7 (4.02) | 4 | 4 (3.60) | 3 | 3 (4.76) |
| Insulin and metformin | 8 | 8 (4.60) | 8 | 8 (7.21) | 0 | 0 (0) |

### Frequency distribution of rs9939609 in the studied population

The frequency distribution of rs9939609 is presented in Table 2. Across the entire population, the minor allele (A) had a prevalence of 17.82%, with 20.72% in the poor control group and 12.70% in the good control group. Notably, the AA homozygous mutated genotype was absent in the good control group. No deviations from HW equilibrium (p > 0.05) were observed in the overall study population or within the good and poor glycemic-control groups.

**Table 2:** Genotype and allele frequencies of rs9939609.

| Genotype and Allele frequencies |  |  |  |  |  |  |  |
| --- | --- | --- | --- | --- | --- | --- | --- |
| Genotype | All | Poor control | Good control | Allele | All | Poor control | Good control |
| TT | 118 (67.82) | 71 (63.97) | 47 (74.60) | T | 286 (82.18) | 176 (79.28) | 110 (87.30) |
| AT | 50 (28.74) | 34 (30.63) | 16 (25.40) | A | 62 (17.82) | 46 (20.72) | 16 (12.70) |
| AA | 6 (3.44) | 6 (5.40) | 0 (0) |  |  |  |  |

### Association between rs9939609-A and glycemic control

Association analyses are presented in Table 3. In the crude (unadjusted) analysis, we observed a statistical trend (p < 0.10) in both the additive and recessive models, suggesting that A-allele carriers may have an increased risk of poor glycemic control. After adjustment for age and sex, both models reached the conventional significance threshold (p < 0.05). In the additive model, each additional copy of the A allele was associated with a 1.15-fold higher likelihood of poor glycemic control. Under the recessive model, individuals homozygous for the A allele (AA) showed a 1.51-fold increased risk of poor glycemic control compared with carriers of the T allele (AT or TT). The association became marginal in both the additive (OR = 1.38; p = 0.059) and recessive (OR = 1.45; p = 0.062) models when further adjusting for years with T2D and treatment. Nevertheless, after further adjustment for BMI, both models strengthened and became statistically significant (p < 0.05), with ORs of 1.15 and 1.51, respectively.

**Table 3:** Association analyses of rs9939609 with glycemic control. Associations with p < 0.05 are highlighted in bold.

| Association analyses with glycemic control |  |  |  |  |  |  |  |  |
| --- | --- | --- | --- | --- | --- | --- | --- | --- |
| Model | Additive<br>OR (IC) | p | Dominant<br>OR (IC) | p | Overdominant<br>OR (IC) | p | Recessive<br>OR (IC) | p |
| Unadjusted | 1.13<br>(0.99-1.29) | 0.063 | 1.12<br>(0.96-1.30) | 0.151 | 1.06<br>(0.91-1.24) | 0.466 | 1.46<br>(0.99-2.15) | 0.061 |
| Age + Sex | 1.15<br>(1.01-1.31) | **0.040** | 1.13<br>(0.97-1.32) | 0.111 | 1.07<br>(0.91-1.25) | 0.414 | 1.49<br>(1.01-2.20) | **0.044** |
| Age + Sex +<br>Treatment +<br>Years with<br>diabetes | 1.38<br>(1.00-1.30) | 0.059 | 1.13<br>(0.96-1.32) | 0.142 | 1.06<br>(0.90-1.25) | 0.461 | 1.45<br>(0.98-2.14) | 0.062 |
| Age + Sex +<br>Treatment +<br>Years with<br>diabetes + BMI | 1.15<br>(1.003-1.31) | **0.047** | 1.13<br>(0.97-1.32) | 0.132 | 1.06<br>(0.90-1.24) | 0.485 | 1.51<br>(1.03-2.23) | **0.038** |

## Discussion

We found an association between rs9939609-A and clinically defined glycemic control. Although a prior study suggested a similar association using simple, unadjusted analyses (30), our work is, to our knowledge, the first to establish this relationship using comprehensive covariate adjustment, including years living with T2D, treatment scheme, and BMI. This substantially increases the robustness of the association. Overall, our findings indicate that carrying at least one copy of the A allele increases the likelihood of poor glycemic control, with an even greater effect observed in homozygous AA individuals, who exhibited up to a 1.51-fold increase in risk.

Adjustment for age, sex, years with T2D, and treatment yielded a marginal association between rs993960-A and glycemic control. However, adding BMI to this model strengthened the effect size and the association became significant. The strengthening of the signal after BMI adjustment indicates a BMI independent genetic effect, consistent with BMI acting as a suppressor variable. This is partly in line with our previous study, in which the A allele of rs9939609 was associated with T2D onset independently of BMI, although a suppressor effect was not observed in that context (23). Moreover, this BMI independent association with T2D has also been reported in Vietnamese (22) and Northern European (21) populations. Taken together, these findings support the notion, previously proposed in our work (23), that *FTO* may influence T2D risk through mechanisms beyond adiposity. The present results suggest that such BMI independent pathways may also operate in the context of glycemic control, warranting further investigation but remaining outside the scope of this article.

Notably, our previous study suggested that rs9939609 does not directly influence fasting glucose in healthy individuals, but may exert its metabolic effects through T2D-related pathways (23). The current finding that the A allele is associated with poorer glycemic control among individuals with T2D is consistent with this interpretation, indicating that the variant’s impact on fasting glucose manifests under conditions of impaired glucose regulation rather than in metabolically healthy states. However, these conclusions remain a potential hypothesis due to the cross-sectional nature of the associations observed previously.

It is important to note that the vast majority of our participants (85%) were receiving oral hypoglycemic agents (OHAs) as their primary treatment regimen. In this context, the observed association between the rs9939609-A allele and poorer glycemic outcomes likely reflects suboptimal glycemic control despite ongoing pharmacologic therapy. This strengthens the relevance of our findings within a treated, real-world population of patients living with T2D. Nonetheless, the present study did not perform stratified or interaction analyses to assess potential differential effects across specific medications, treatment combinations, or insulin use, as the sample size limited the power of such analyses. Future studies with larger cohorts and richer pharmacologic granularity are needed to determine whether the rs9939609-A allele confers pharmacogenomic effects with respect to particular glucose-lowering agents or treatment schemes.

Another important characteristic of our study is that the participants were recruited in Yucatan, Mexico, a region in Southeastern Mexico where Mayan ancestry is prevalent and the population is admixed (31,32). A limitation is that we did not measure ancestry informative markers to confirm individual ancestry proportions. However, given that our previous study showed that the A allele of rs9939609 was associated with T2D, in a manner consistent with findings from the majority of global studies (23), and that in the present study the same A allele is associated with poorer glycemic control, it is unlikely that ancestry represents a major confounder in our analysis, although future studies incorporating ancestry informative markers would help confirm this.

Strengths of this study include a well characterized cohort with key variables available for modeling, including specific treatment regimens and years with T2D, which could be appropriately accounted for in the regression analyses. However, despite having treatment information, the sample size limited our ability to conduct stratified or interaction analyses to determine whether particular medications or combinations of therapies drive the association between rs9939609-A and glycemic control. Additionally, we did not include ancestry informative markers to quantify individual ancestry; nevertheless, the risk direction for the A allele aligns with the majority of reports on *FTO* and cardiometabolic outcomes, which provides some reassurance regarding confounding by ancestry. Finally, the moderate to small sample size may lead to imprecision in effect estimates. Therefore, replication is needed to validate the associations reported in this paper.

In conclusion, rs9939609-A may be associated with poor glycemic control. These results also raise the possibility that other variants previously identified for T2D susceptibility could have a role in glycemic control. Future studies should assess the generalizability of these findings across diverse ancestries using independent cohorts and cross-ancestry meta-analyses. Furthermore, larger studies incorporating medication and treatment stratification would be necessary to test potential fine-grained pharmacogenomic hypotheses.

## Data availability

Data will be provided upon reasonable request from the corresponding author to safeguard patient anonymity and confidentiality.

## Acknowledgments

We thank Professor. Maria Guadalupe García Escalante for historically being a vital part for the project foundation and data collection. We are very thankful to all participants who were part of this study. Special thanks to the thesis, social service and internship students for their incredible valuable contributions throughout the project.

## Contributions

N.F.-B and N.V.-G. conceptualized and designed the study. N.V.-G collected the data and co-acquired the funding. N.F.-B. performed the core statistical analyses and wrote the manuscript. R.V.E.-C contributed to the genotyping assays, laboratory work and data curation. I.Q.-O. L.V.-G and G.V.-P supported sample processing, laboratory assessments, and provided the necessary infrastructure for these procedures. N.F.-B. and N.V.-G. share senior authorship for this work.

## References

1. Kasuga M. Insulin resistance and pancreatic β cell failure. J Clin Invest. 2006 Jul 3;116(7):1756–60. doi:10.1172/JCI29189

2. Duncan BB, Magliano DJ, Boyko EJ. IDF Diabetes Atlas 11th edition 2025: global prevalence and projections for 2050. Nephrol Dial Transplant. 2026 Jan 1;41(1):7–9. doi:10.1093/ndt/gfaf177

3. Ganesan K, Rana MBM, Sultan S. Oral Hypoglycemic Medications. In: StatPearls. Treasure Island (FL): StatPearls Publishing; 2025. PubMed PMID: 29494008.

4. Stolar MW. Defining and achieving treatment success in patients with type 2 diabetes mellitus. Mayo Clin Proc. 2010 Dec;85(12 Suppl):S50–59. doi:10.4065/mcp.2010.0471 PubMed PMID: 21106864; PubMed Central PMCID: PMC2996162.

5. American Diabetes Association Professional Practice Committee for Diabetes*. 6. Glycemic Goals, Hypoglycemia, and Hyperglycemic Crises: Standards of Care in Diabetes—2026. Diabetes Care. 2025 Dec 8;49(Supplement_1):S132–49. doi:10.2337/dc26-S006

6. Goudswaard AN, Furlong NJ, Rutten GEHM, Stolk RP, Valk GD. Insulin monotherapy versus combinations of insulin with oral hypoglycaemic agents in patients with type 2 diabetes mellitus. Cochrane Database Syst Rev. 2004 Oct 18;2004(4):CD003418. doi:10.1002/14651858.CD003418.pub2 PubMed PMID: 15495054; PubMed Central PMCID: PMC9007040.

7. Tarekegn ET, Gobezie MY, Haile MB, Zerga AA. Glycemic control and associated factors among type 2 diabetes patients attending at Dessie comprehensive specialized hospital outpatient department. Sci Rep. 2025 Mar 18;15(1):9286. doi:10.1038/s41598-025-93739-2

8. van Leeuwen N, Nijpels G, Becker ML, Deshmukh H, Zhou K, Stricker BHC, et al. A gene variant near ATM is significantly associated with metformin treatment response in type 2 diabetes: a replication and meta-analysis of five cohorts. Diabetologia. 2012 Jul 1;55(7):1971–7. doi:10.1007/s00125-012-2537-x

9. Dawed AY, Yee SW, Zhou K, van Leeuwen N, Zhang Y, Siddiqui MK, et al. Genome-Wide Meta-analysis Identifies Genetic Variants Associated With Glycemic Response to Sulfonylureas. Diabetes Care. 2021 Oct 4;44(12):2673–82. doi:10.2337/dc21-1152

10. Li JH, Brenner LN, Kaur V, Figueroa K, Schroeder P, Huerta-Chagoya A, et al. Genome-wide association analysis identifies ancestry-specific genetic variation associated with acute response to metformin and glipizide in SUGAR-MGH. Diabetologia. 2023 Jul 1;66(7):1260–72. doi:10.1007/s00125-023-05922-7

11. Wu B, Yee SW, Xiao S, Xu F, Sridhar SB, Yang M, et al. Genome-Wide Association Study Identifies Pharmacogenomic Variants Associated With Metformin Glycemic Response in African American Patients With Type 2 Diabetes. Diabetes Care. 2023 Aug 28;47(2):208–15. doi:10.2337/dc22-2494

12. Zhou K, Yee SW, Seiser EL, van Leeuwen N, Tavendale R, Bennett AJ, et al. Variation in the glucose transporter gene SLC2A2 is associated with glycemic response to metformin. Nat Genet. 2016 Sep 1;48(9):1055–9. doi:10.1038/ng.3632

13. Rotroff DM, Yee SW, Zhou K, Marvel SW, Shah HS, Jack JR, et al. Genetic Variants in CPA6 and PRPF31 Are Associated With Variation in Response to Metformin in Individuals With Type 2 Diabetes. Diabetes. 2018 Apr 12;67(7):1428–40. doi:10.2337/db17-1164

14. Li JH, Perry JA, Jablonski KA, Srinivasan S, Chen L, Todd JN, et al. Identification of Genetic Variation Influencing Metformin Response in a Multiancestry Genome-Wide Association Study in the Diabetes Prevention Program (DPP). Diabetes. 2023 Dec 16;72(8):1161–72. doi:10.2337/db22-0702

15. Zhou K, Bellenguez C, Spencer CCA, Bennett AJ, Coleman RL, Tavendale R, et al. Common variants near ATM are associated with glycemic response to metformin in type 2 diabetes. Nat Genet. 2011 Feb 1;43(2):117–20. doi:10.1038/ng.735

16. Suzuki K, Hatzikotoulas K, Southam L, Taylor HJ, Yin X, Lorenz KM, et al. Genetic drivers of heterogeneity in type 2 diabetes pathophysiology. Nature. 2024 Mar 1;627(8003):347–57. doi:10.1038/s41586-024-07019-6

17. Popović AM, Huđek Turković A, Žuna K, Bačun-Družina V, Rubelj I, Matovinović M. Gene Polymorphisms at the Crossroads of Metabolic Pathways of Obesity and Epigenetic Influences. Food Technol Biotechnol. 2023;61(1):14–26. doi:10.17113/ftb.61.01.23.7594

18. Ali AHA, Shkurat TP, Abbas AH. Association analysis of FTO gene polymorphisms rs9939609 and obesity risk among the adults: A systematic review and meta-analysis. Meta Gene. 2021 Feb 1;27:100832. doi:10.1016/j.mgene.2020.100832

19. Frayling TM, Timpson NJ, Weedon MN, Zeggini E, Freathy RM, Lindgren CM, et al. A Common Variant in the FTO Gene Is Associated with Body Mass Index and Predisposes to Childhood and Adult Obesity. Science. 2007 May 11;316(5826):889–94. doi:10.1126/science.1141634

20. Amine Ikhanjal M, Ali Elouarid M, Zouine C, El alami H, Errafii K, Ghazal H, et al. FTO gene variants (rs9939609, rs8050136 and rs17817449) and type 2 diabetes mellitus risk: A Meta-Analysis. Gene. 2023 Dec 15;887:147791. doi:10.1016/j.gene.2023.147791

21. Hertel JK, Johansson S, Sonestedt E, Jonsson A, Lie RT, Platou CGP, et al. FTO, Type 2 Diabetes, and Weight Gain Throughout Adult Life: A Meta-Analysis of 41,504 Subjects From the Scandinavian HUNT, MDC, and MPP Studies. Diabetes. 2011 Apr 23;60(5):1637–44. doi:10.2337/db10-1340

22. Binh TQ, Phuong PT, Nhung BT, Thoang DD, Lien HT, Thanh DV. Association of the common FTO-rs9939609 polymorphism with type 2 diabetes, independent of obesity-related traits in a Vietnamese population. Gene. 2013 Jan 15;513(1):31–5. doi:10.1016/j.gene.2012.10.082

23. Fragoso-Bargas N, Toloza-Couoh LN, Quintal-Ortiz I, Valencia-Pacheco G, Valadez-Gonzalez N. Exploring the Association of FTO rs9939609 with Type 2 Diabetes, Fasting Glucose and HbA1c in a Southeastern Mexican Region of Predominant Mayan Genetic Background. Biomolecules. 2025;15(11):1492. doi:10.3390/biom15111492

24. Pinto-Escalante D del C, Ruiz-García L, García-Escalante MG, Valadez-González N, Rubi-Castellanos R, Vera-Gamboa L, et al. Análisis genealógico de diabetes tipo 2 en familias extensas multigeneracionales de una comunidad yucateca. Rev Bioméd Vol 35 Núm 1 2024. 2023. doi:10.32776/revbiomed.v35i1.1136

25. Wigginton JE, Cutler DJ, Abecasis GR. A Note on Exact Tests of Hardy-Weinberg Equilibrium. Am J Hum Genet. 2005 May 1;76(5):887–93. doi:10.1086/429864

26. Graffelman J, Camarena JM. Graphical Tests for Hardy-Weinberg Equilibrium Based on the Ternary Plot. Hum Hered. 2007 Sep 26;65(2):77–84. doi:10.1159/000108939

27. Graffelman J. Exploring Diallelic Genetic Markers: The HardyWeinberg Package. J Stat Softw. 2015 Mar 20;64(3):1–23. doi:10.18637/jss.v064.i03

28. Graffelman J, Weir BS. Testing for Hardy–Weinberg equilibrium at biallelic genetic markers on the X chromosome. Heredity. 2016 Jun 1;116(6):558–68. doi:10.1038/hdy.2016.20

29. Mahajan A, Spracklen CN, Zhang W, Ng MCY, Petty LE, Kitajima H, et al. Multi-ancestry genetic study of type 2 diabetes highlights the power of diverse populations for discovery and translation. Nat Genet. 2022 May 1;54(5):560–72. doi:10.1038/s41588-022-01058-3

30. Hussain M, Waheed A, Elahi A, Mustafa G. Fat Mass and Obesity-Related (FTO) Gene Variant Is a Predictor of CVD in T2DM Patients. J Diabetes Res. 2024 Jan 1;2024(1):5914316. doi:10.1155/2024/5914316

31. González-Herrera L, Sosa-Escalante JE, López-González P, López-González MJ, Gamboa-Magaña RY, Herrera-Diaz RG, et al. Ancestral proportions based on 22 autosomal STRs of an admixed population (Mestizos) from the Península of Yucatán, México. Forensic Sci Int Genet Suppl Ser. 2019 Dec 1;7(1):429–31. doi:10.1016/j.fsigss.2019.10.039

32. Silva-Zolezzi I, Hidalgo-Miranda A, Estrada-Gil J, Fernandez-Lopez JC, Uribe-Figueroa L, Contreras A, et al. Analysis of genomic diversity in Mexican Mestizo populations to develop genomic medicine in Mexico. Proc Natl Acad Sci. 2009 May 26;106(21):8611–6. doi:10.1073/pnas.0903045106

